# The Association of Serum Lipid and Lipoprotein Levels with Total and Differential Leukocyte Counts: Results of a Cross-sectional and Longitudinal Analysis of the UK Biobank

**DOI:** 10.1101/2020.07.11.20149310

**Authors:** Bradley Tucker, Sonia Sawant, Hannah McDonald, Kerry-Anne Rye, Sanjay Patel, Kwok Leung Ong, Blake J Cochran

**Affiliations:** Heart Research Institute, Sydney, Australia; Faculty of Medicine and Health, University of Sydney, Sydney, Australia; School of Medical Sciences, UNSW, Sydney, Australia; Royal Prince Alfred Hospital, Sydney, Australia; Bankstown-Lidcombe Hospital, Sydney, Australia

**Keywords:** Inflammation, Lipids and lipoproteins, Cholesterol, Myeloid cells, Atherosclerosis

## Abstract

**Background and aims:** There is some evidence of a cross-sectional, and possibly causal, relationship of lipid levels with leukocyte counts in mice and humans. This study investigates the cross-sectional and longitudinal relationship of blood lipid and lipoprotein levels with leukocyte counts in the UK Biobank cohort.

**Methods:** The primary cross-sectional analysis included 417,132 participants with valid data on lipid measures and leukocyte counts. A subgroup analysis was performed in 333,668 participants with valid data on lipoprotein(a). The longitudinal analysis included 9,058 participants with valid baseline and follow-up data on lipid and lipoprotein levels and leukocyte counts. The association of lipid and lipoprotein levels with leukocyte counts was analysed by multivariable linear regression.

**Results:** Several relationships were significant in both cross-sectional and longitudinal analysis. After adjustment for demographic, socioeconomic and other confounding factors a higher eosinophil count was associated with lower HDL cholesterol and apolipoproteinA-I concentration (p<0.001). Higher triglycerides levels were associated with higher total leukocyte, basophil, eosinophil, monocyte and neutrophil counts (all p<0.01). A higher lymphocyte count was associated with a higher apolipoprotein B level (p<0.001). In the longitudinal analysis lipoprotein(a) was inversely associated with basophil count in men but not women (p<0.001).

**Conclusion:** Triglyceride levels demonstrate a robust positive association with total and differential leukocyte counts suggesting they may be directly involved in leuokogenesis. However, unlike in murine models, the remainder of these relationships are modest which suggests that cholesterol and lipoproteins are minimally involved in leukogenesis in humans.

## Introduction

Atherosclerosis is a chronic inflammatory disease, characterised by lipid and leukocyte accumulation within the intima of medium and large-sized arteries^1^. Over time, atherosclerotic lesions can cause ischemia by significantly reducing blood supply to downstream tissues. Highly inflamed plaques are also prone to rupture and are a major precipitating factor in myocardial infarction and stroke^2^.

There is an abundance of literature highlighting the role of various lipids and lipoproteins in atherosclerosis^3, 4^. Elevated serum levels of low-density lipoprotein (LDL) cholesterol, triglycerides, apolipoprotein B (apoB) and lipoprotein(a) [Lp(a)] are known to be independently associated with increased cardiac events^5-7^. On the other hand, elevated levels of high-density lipoprotein (HDL) cholesterol and apolipoprotein A-I (apoA-I) are associated with a reduced risk of cardiovascular disease (CVD)^3^. Similarly, preclinical and clinical studies have highlighted the role of specific immune cells in the development and progression of atherosclerosis^1, 2, 8^. Elevated total leukocyte count has been shown to predict CVD^9^. However, the precise interaction between leukocyte counts and CVD remains ill-defined.

Results of preclinical studies suggest that lipids are directly involved in leukogenesis^10^. Hypercholesterolemia in mice induces monocytosis, neutrophilia and mobilisation of hematopoietic stem cells (HSC) from the bone marrow into the peripheral circulation, likely via disruption of the stromal cell-derived factor-1:chemokine receptor 4 axis^11-13^. Despite this evidence, there is limited data to indicate a similar phenomenon in humans. Liu *et al*.^14^ reported a positive association of total leukocyte count with total cholesterol, LDL cholesterol and triglycerides. A recent analysis of the National Health and Nutrition Examination Survey (NHANES)^15^ demonstrated a strong positive correlation of triglycerides with both lymphocyte and basophil counts. Results from the Multi-Ethnic Study of Atherosclerosis (MESA) showed that total cholesterol, LDL cholesterol and HDL cholesterol were inversely associated with total leukocyte count; whereas, triglycerides were positively associated with total leukocyte count^16^. Results from the NHANES^15^ indicated that the effect of lipids on leukocyte counts was sex dependent, although this was not supported by findings from the MESA^16^.

The present study provides a more robust estimate of the cross-sectional relationship of lipid levels with differential leukocyte counts utilising a large cohort from the UK Biobank. Furthermore, it investigates the longitudinal relationship of changes in serum lipids and lipoproteins with changes in total and differential leukocyte counts. We also assess whether sex differentially influences any of these relationships.

## Methods

### Participant selection

The UK Biobank is a large prospective study of ∼500,000 participants aged 37-73 years from the general population of the United Kingdom, recruited between 2007 and 2010. Participants attended one of 22 recruitment centres where they underwent baseline assessment. A sub-sample of study participants (n∼20,000) underwent repeat examination between 2010 and 2013.

Among 502,524 participants, 477,303 had valid data on total and differential leukocyte counts at baseline, with 417,132 of these having valid data on total cholesterol, HDL cholesterol, LDL cholesterol and triglycerides (Figure 1). As a large number of participants had missing data on Lp(a) a subgroup analysis was performed on those with valid Lp(a) data (*n=*333,668). Follow-up data was available for 9,058 of the participants included in the primary analysis.

**Figure 1:**
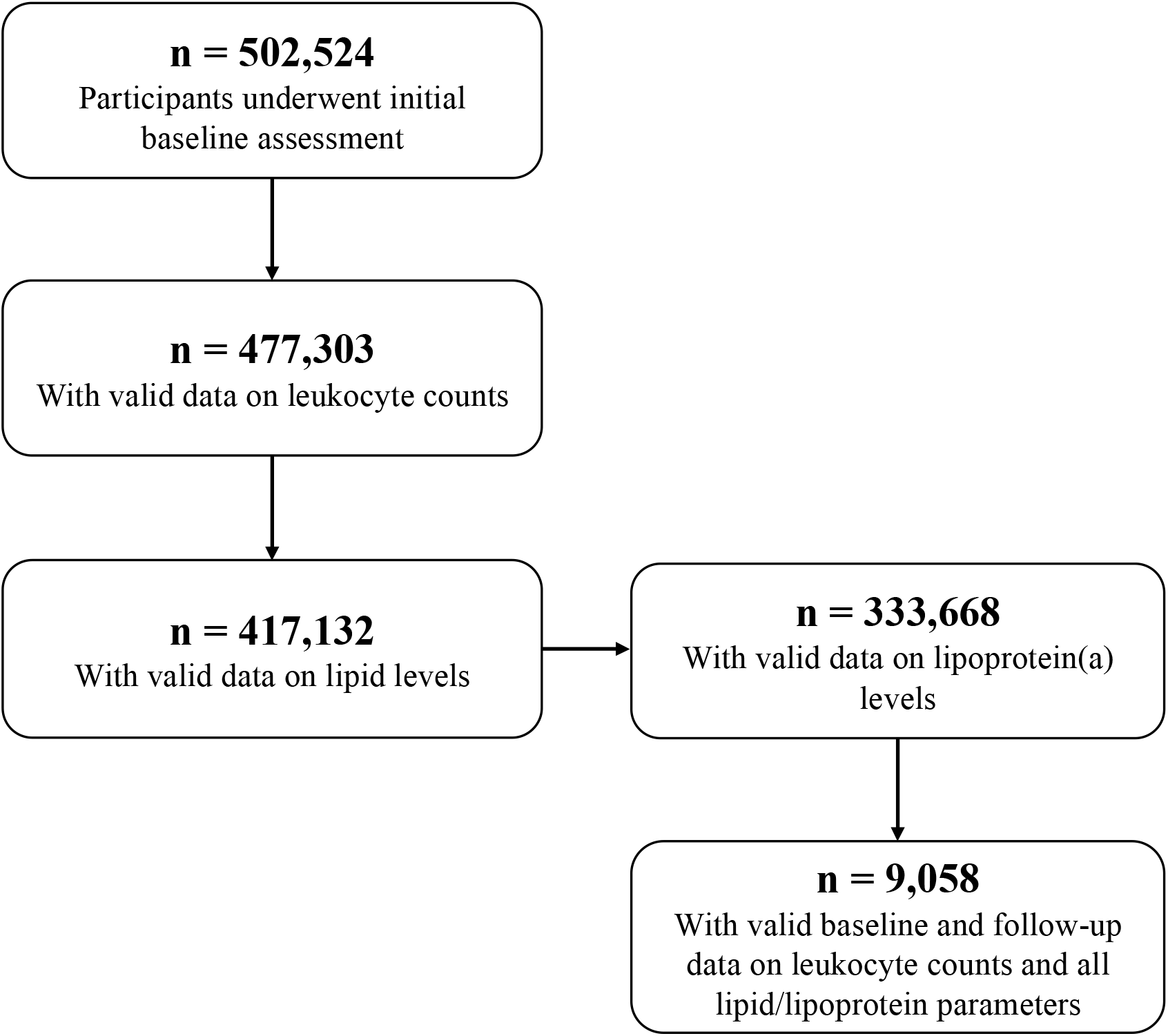
Participant flow. Of the 502,524 participants included at baseline, 417,132 participants had valid data on leukocyte counts and lipid levels. Of these, a sub-cohort of 333,668 participants had Lp(a) levels measured and 9,058 had valid follow-up data.

### Leukocyte count and lipid level measurement

The UK Biobank sample handling procedures have been previously outlined^17^. Non-fasting blood samples were collected from participants and cell counts were obtained within 24 h using Beckman Coulter LH750 haematology analysers at the UK Biobank Central Laboratory (Manchester, United Kingdom). All lipid and lipoprotein levels were measured directly on a Beckman Coulter AU5800 chemistry analyser.

### Other covariates of interest

At baseline examination, demographics, socioeconomic status, medication usage, alcohol intake, smoking status, pack-years smoking, and medical history including CVD and diabetes were obtained via self-reported questionnaire^18^. The Townsend deprivation index (a measure of socio-economic status) for each participant was calculated at recruitment, based on postcode^19^. History of CVD was defined as any self-reported myocardial infarction, angina or stroke prior to the date of recruitment. Height and weight were recorded to calculate body mass index (BMI). The average of two separate blood pressure readings was used as the reported value. Serum creatinine, high-sensitivity C-reactive protein (CRP) and urea were measured on a Beckman Coulter AU5800 chemistry analyzer. Glycated haemoglobin (HbA1c) was measured in serum by high-performance liquid chromatography on a Bio-Rad VARIANT II Turbo.

### Statistical analysis

SPSS 24 (IBM, Armonk, NY) and STATA 14.0 (StataCorp, College Station, TX) were used for statistical analysis. Variables with a skewed distribution are presented as median (interquartile range) and subjected to natural logarithm (ln) transformation prior to analysis to improve normality. For the basophil count, a large number of participants had values of zero. In these cases, a constant of 1 was added to the basophil count before ln-transformation. For the Townsend deprivation index, data were highly skewed and included a large proportion of negative values. Negative values were addressed by adding the constant 7.26 (as the lowest value was 6.26) to each individual Townsend deprivation index prior to ln-transformation, so that min(*x* + a) = 1; where *x* = Townsend deprivation index and a = constant. As Lp(a) levels were measured in a sub-cohort of 333,668 participants, analysis for the relationship of Lp(a) levels with leukocyte counts was conducted in this sub-cohort.

For the cross-sectional analysis, multivariable linear regression analysis was performed using robust standard error estimation. The regression coefficient (B) was estimated as the ln-transformed cell counts per one standard deviation (SD) increase in lipid levels. In model 1, data were adjusted for demographic factors including age, sex, and Townsend deprivation index. In model 2, data were further adjusted for CVD risk factors including BMI, smoking status, pack-years of smoking, alcohol consumption status, systolic blood pressure, use of anti-hypertensive medication, diabetes, HbA1c, serum urea, serum creatinine, CRP, use of lipid-lowering medication and history of CVD. In model 3, data were further adjusted for total cholesterol, HDL cholesterol, LDL cholesterol, triglycerides, apoA-I and apoB, where appropriate. In model 4, data were further adjusted for red blood cell and platelet counts. The regression coefficient was converted to a percentage change using the following formula: % change = e^(*B*-1)^ × 100.

For the longitudinal analysis, absolute annual change in leukocyte counts and lipid levels were included in the final regression model as dependent and independent variables, respectively. Data were adjusted for the same set of covariates as the cross-sectional analysis, with further adjustment for the use of lipid-lowering medications at the repeat assessment (Model 4) and baseline levels of the leukocyte count and lipid parameter of interest (Model 5).

In all of the analyses, no multicollinearity issue was detected (all variance inflation factors <4.0). Multiple testing correction of 35 tests (7 lipid traits x 5 leukocyte parameters) was performed using false discovery rate with the study-wide false discovery rate at 0.05. The interaction of sex was estimated by including the interaction term in the regression model in the full sample after adjusting for the main effects of the covariates. In cases where a significant interaction was detected, results were stratified by sex. A two-tailed *p*<0.05 was considered statistically significant.

## Results

### Baseline characteristics

Table 1 shows the baseline characteristics of participants included in the cross-sectional and longitudinal analyses. For participants in the cross-sectional analysis the mean age was 56.6 years, 53.7% were female, 10.6% were current smokers and 92.0% consumed alcohol. The mean total cholesterol in these participants was 5.69 mmol/L and the median total leukocyte count was 6.66×10^9^ cells/L. For participants in the longitudinal analysis the mean age was 57.3 years, 48.6% were female, 6.6% were current smokers and 94.6% consumed alcohol. The mean total cholesterol in this population was 5.65 mmol/L and the median total leukocyte count was 6.41×10^9^ cells/L.

**Table 1:** Baseline characteristic of study participants.

|  | **Cross-sectional analysis** |  | **Longitudinal analysis** |  |
| --- | --- | --- | --- | --- |
| **Characteristic** | **n** | **Count** | **n** | **Count** |
| Age (years), mean (SD) | 417,132 | 56.6 (8.1) | 9,058 | 57.3 (7.4) |
| Female, n (%) | 417,132 | 224,105 (53.7) | 9,058 | 4402 (48.6) |
| Townsend deprivation index, median (IQR) | 416,606 | -2.1 (-3.7, 0.5) | 9,052 | -2.7 (-4.0, -0.8) |
| **Smoking status, n (%)** | 415,027 |  | 9,032 |  |
| Never |  | 226,746 (54.6) |  | 5387 (59.6) |
| Former |  | 144,341 (34.8) |  | 3050 (33.8) |
| Current |  | 43,940 (10.6) |  | 595 (6.6) |
| Pack years of smoking, mean (SD) | 415,027 | 7.1 (14.9) | 9,032 | 5.4 (12.8) |
| **Alcohol drinker status, n (%)** | 416,093 |  | 9,056 |  |
| Never |  | 18,273 (4.4) |  | 263 (2.9) |
| Former |  | 14,980 (3.6) |  | 230 (2.5) |
| Current |  | 382,840 (92.0) |  | 8563 (94.6) |
| BMI (kg/m<sup>2</sup>), mean (SD) | 415,471 | 27.4 (4.8) | 9,036 | 26.8 (4.4) |
| Systolic blood pressure (mmHg), mean (SD) | 416,653 | 139.8 (19.7) | 9,049 | 139.5 (18.9) |
| Lipid-lowering medication, n (%) | 413,527 | 72,940 (17.6) | 9,020 | 1430 (15.9) |
| Blood pressure medication, n (%) | 413,527 | 87,155 (21.1) | 9,020 | 1623 (18.0) |
| Diagnosis of diabetes, n (%) | 415,315 | 21,688 (5.2) | 9,044 | 349 (3.9) |
| Creatinine (μmol/L), median (IQR) | 417,083 | 70.5 (61.4, 81.0) | 9,057 | 71.6 (62.3, 82.0) |
| Urea (mmol/L), median (IQR) | 416,986 | 5.27 (4.49, 6.15) | 9,056 | 5.33 (4.57, 6.16) |
| CRP (mg/L), median (IQR) | 416,232 | 1.34 (0.66, 2.78) | 9,040 | 1.17 (0.59, 2.34) |
| HbA1c (%), median (IQR) | 395,960 | 35.2 (32.8, 37.9) | 8,550 | 35.0 (32.6, 37.4) |
| History of CVD, n (%) | 415,657 | 24,072 (5.8) | 9,044 | 396 (4.4) |
| **Lipid profile, mean (SD) or median (IQR) where appropriate** |  |  |  |  |
| Total cholesterol (mmol/L) | 417,132 | 5.69 (1.14) | 9,058 | 5.65 (1.10) |
| HDL cholesterol (mmol/L) | 417,132 | 1.45 (0.38) | 9,058 | 1.44 (0.36) |
| LDL cholesterol (mmol/L) | 417,132 | 3.56 (0.87) | 9,058 | 3.54 (0.84) |
| Triglycerides (mmol/L) | 417,132 | 1.48 (1.05, 2.15) | 9,058 | 1.45 (1.03, 2.09) |
| apoA-I (g/L) | 414,685 | 1.54 (0.27) | 9,058 | 1.53 (0.26) |
| apoB (g/L) | 415,044 | 1.03 (0.24) | 9,058 | 1.02 (0.23) |
| Lipoprotein A (nmol/L) | 333,668 | 21.1 (9.6, 62.1) | 9,058 | 20.9 (9.7, 57.8) |
| **Blood cell counts, mean (SD) or median (IQR) where appropriate** |  |  |  |  |
| Basophils ($\times 10^9$ cells/L) | 417,132 | 0.02 (0.00, 0.04) | 9,058 | 0.02 (0.00, 0.04) |
| Eosinophils ($\times 10^9$ cells/L) | 417,132 | 0.14 (0.10, 0.21) | 9,058 | 0.13 (0.10, 0.20) |
| Lymphocytes ($\times 10^9$ cells/L) | 417,132 | 1.88 (1.50, 2.30) | 9,058 | 1.80 (1.50, 2.20) |
| Monocytes ($\times 10^9$ cells/L) | 417,132 | 0.45 (0.36, 0.57) | 9,058 | 0.45 (0.37, 0.56) |
| Neutrophils ($\times 10^9$ cells/L) | 417,132 | 4.02 (3.27, 4.97) | 9,058 | 3.90 (3.20, 4.71) |
| Total leukocytes ($\times 10^9$ cells/L) | 417,132 | 6.66 (5.63, 7.86) | 9,058 | 6.41 (5.50, 7.50) |
| RBCs ($\times 10^{12}$ cells/L) | 417,131 | 4.52 (0.42) | 9,058 | 4.53 (0.40) |
| Platelets ($\times 10^9$ cells/L) | 417,128 | 248.0 (213.2, 287.0) | 9,058 | 245.3 (211.2, 282.9) |

Data are expressed as mean (standard deviation), number (percentage) or median (interquartile range), where appropriate.
**Abbreviations:** apoA-I, apolipoprotein A-I; apoB, apolipoprotein B; BMI, body mass index; CRP, C-reactive protein; CVD, cardiovascular disease; HbA1c, glycated haemoglobin A1c; HDL, high-density lipoprotein; LDL, low-density lipoprotein; RBC, red blood cell

Participants excluded from the cross-sectional analysis were more likely to be younger and female (Supplementary Table 1). Those with Lp(a) measurement were more likely to be younger, female and a never smoker than those without. Finally, compared to those in the cross-sectional analysis those participants included in the longitudinal analysis were more likely to be older, male, have a lower BMI and be a non-smoker. However, all these differences were very modest.

### Association of total and LDL cholesterol with leukocyte counts

In the cross-sectional and longitudinal analyses, results for total and LDL cholesterol were largely equivalent. In the cross-sectional analysis (Figure 2, Supplementary Table 2) total and LDL cholesterol was inversely associated with total leukocyte, basophil, eosinophil, monocyte and neutrophil counts (all p<0.001). Each one-SD increment in total cholesterol (1.14 mmol/L) was associated with 2.7%, 0.1%, 2.0%, 3.3% and 3.7% lower total leukocyte, basophil, eosinophil, monocyte and neutrophil counts respectively. Each one-SD increment in LDL cholesterol (0.87 mmol/L) was associated with 2.4%, 0.1%, 1.7%, 3.1% and 3.3% lower total leukocyte, basophil, eosinophil, monocyte and neutrophil counts respectively. An interaction with sex was detected in the association of total and LDL cholesterol with total leukocyte, eosinophil, monocyte and neutrophil counts (all p<0.05); whereby the associations were more prominent in women than men (Supplementary Table 3). There was no evidence of a relationship between LDL cholesterol and lymphocyte count in the full sample. However, an interaction with sex was detected (p=0.003) which revealed a positive association in men yet an inverse association in women (Supplementary Table 3).

**Figure 2:**
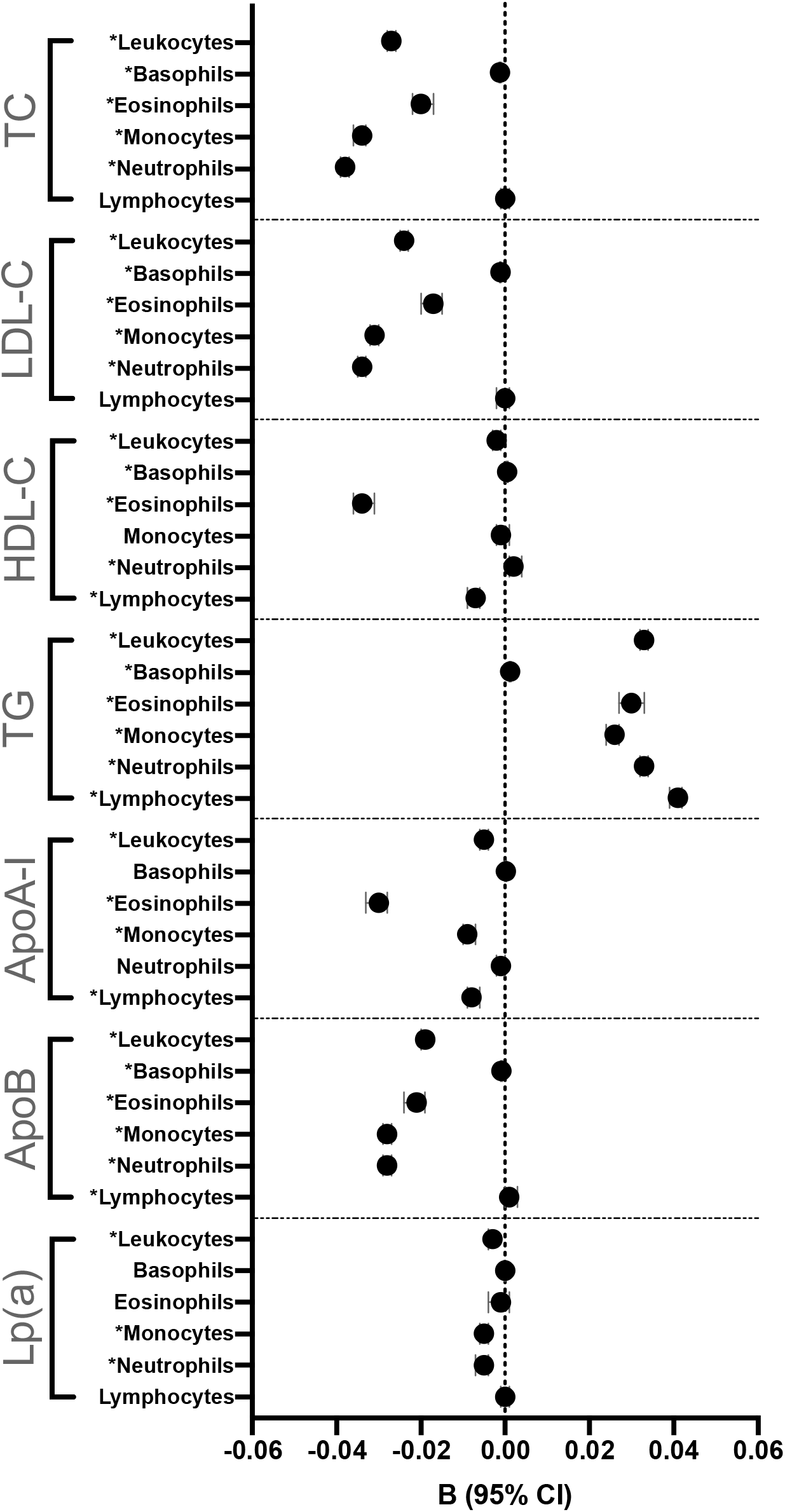
Cross-sectional analysis of the relationship of lipid and lipoprotein levels with WBC counts. Regression coefficient B (95% confidence interval [CI]) was expressed as ln-transformed cell counts (x10^9^ cells/L) per one-SD unit increase in lipid or lipoprotein level. Data on triglycerides and Lp(a) were ln-transformed in the analysis. Data were adjusted for age, sex, Townsend deprivation index, BMI, smoking status, pack-years of smoking, alcohol drinker status, systolic blood pressure, use of anti-hypertensive medication, diabetes, ln-transformed HbA1c, ln-transformed serum urea, ln-transformed serum creatinine, ln-transformed CRP, use of lipid-lowering medication, history of CVD, RBC count and platelet count. Date were further adjusted for: HDL cholesterol and ln-transformed triglycerides for the analysis of total cholesterol and LDL cholesterol; LDL cholesterol and ln-transformed triglycerides for the analysis of HDL cholesterol; HDL and LDL cholesterol for the analysis of ln-transformed triglycerides and ln-transformed Lp(a); apoB and ln-transformed triglycerides for the analysis of apoA-I; and apoA-I and ln-transformed triglycerides for the analysis of Lp(a). Asterisks (*) indicate p-values that remained significant after correcting for multiple comparisons. **Abbreviations**: apoA-I, apolipoprotein A-I; apoB, apolipoprotein B; BMI, body mass index; CRP, C-reactive protein; HbA1c, glycated haemoglobin; HDL-C, high-density lipoprotein cholesterol; LDL-C, low-density lipoprotein cholesterol; Lp(a), lipoprotein(a); TC, total cholesterol; TGs, triglycerides; RBC, red blood cell.

In the longitudinal analysis (Figure 3, Supplementary Table 4), the average change in total and LDL cholesterol over the mean follow up period of 3.8 years was 0.03±0.93 and 0.00±0.72 mmol/L respectively. In the full adjustment model, there was a significant positive relationship of change in total and LDL cholesterol with change in lymphocyte count (both p<0.001). An annual increase of 1 mmol/L in total and LDL cholesterol was associated with an annual increase in lymphocyte count of 0.096×10^9^ and 0.129×10^9^ cells/L respectively. Change in total but not LDL cholesterol was positively associated with change in total leukocyte count (p=0.003) with an annual increase of 1 mmol/L in total cholesterol being associated with an annual increase in total leukocyte count of 0.102×10^9^ cells/L.

**Figure 3:**
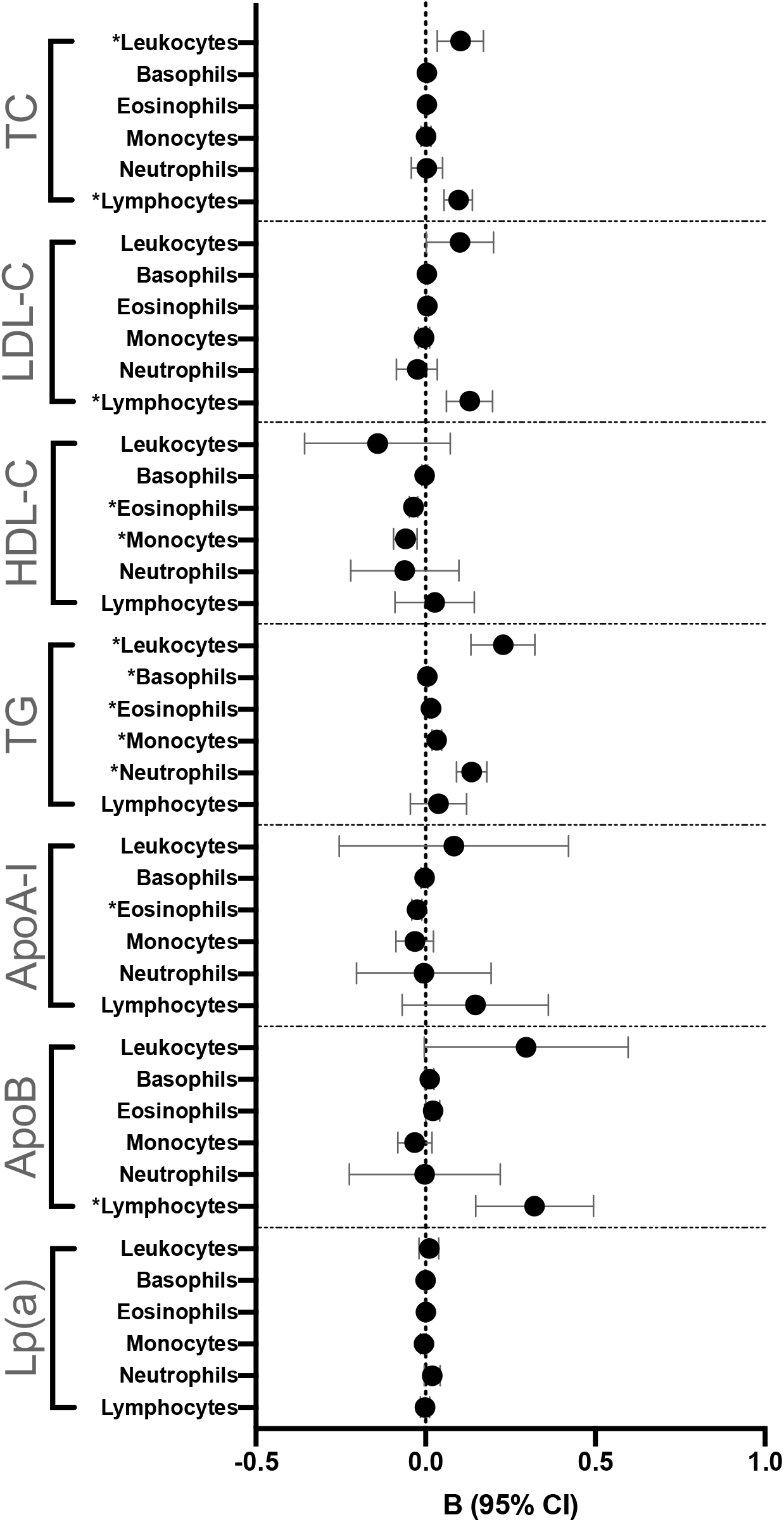
Longitudinal analysis of the relationship of lipid and lipoprotein levels with WBC counts (n=9058). Regression coefficient B (95% confidence interval [CI]) is expressed as annual absolute change in cell counts (x10^9^ cells/L) per one mmol/L increase in total cholesterol, HDL cholesterol, LDL cholesterol and triglycerides; per 1 g/L of apoA-I and apoB; and per 10 nmol/L of Lp(a) per year of follow up. Data were adjusted for age, sex, Townsend deprivation index, BMI, smoking status, pack-years of smoking, alcohol drinker status, systolic blood pressure, use of anti-hypertensive medication, diabetes, ln-transformed HbA1c, ln-transformed serum urea, ln-transformed serum creatinine, ln-transformed CRP, use of lipid-lowering medication at baseline, use of lipid-lowering medication at repeat assessment, history of CVD, RBC count and platelet count. Date were further adjusted for: HDL cholesterol and ln-transformed triglycerides for the analysis of total cholesterol and LDL cholesterol; LDL cholesterol and ln-transformed triglycerides for the analysis of HDL cholesterol; HDL and LDL cholesterol for the analysis of ln-transformed triglycerides and ln-transformed Lp(a); apoB and ln-transformed triglycerides for the analysis of apoA-I; and apoA-I and ln-transformed triglycerides for the analysis of Lp(a). Data were further adjusted for baseline levels of the lipid parameter and WBC count of interest. Asterisks (*) indicate p-values that remained significant after correcting for multiple comparisons. **Abbreviations**: apoA-I, apolipoprotein A-I; apoB, apolipoprotein B; BMI, body mass index; CRP, C-reactive protein; HbA1c, glycated haemoglobin; HDL-C, high-density lipoprotein cholesterol; LDL-C, low-density lipoprotein cholesterol; Lp(a), lipoprotein(a); TC, total cholesterol; TGs, triglycerides; RBC, red blood cell.

### Association of HDL cholesterol with leukocyte counts

In the cross-sectional analysis (Figure 2, Supplementary Table 2), there was a significant inverse association of HDL cholesterol with total leukocyte, eosinophil and lymphocyte counts (all p<0.001). Each one-SD increment in HDL cholesterol (0.38 mmol/L) was associated with 0.2%, 3.3% and 0.7% lower total leukocyte, eosinophil and lymphocyte counts, respectively. In contrast, HDL cholesterol was positively associated with basophil and neutrophil counts (p<0.001 and p=0.001). Each one-SD increment in HDL cholesterol was associated with 0.05% and 0.2% higher basophil and neutrophil counts, respectively. A significant interaction with sex was found in the association of HDL cholesterol with total leukocyte, basophil, neutrophil and lymphocyte counts (all p<0.001), with the relationship being more pronounced in men compared to women (Supplementary Table 3).

In the longitudinal analysis (Figure 3, Supplementary Table 4), the average change in HDL cholesterol over the follow up period was 0.07±0.21 mmol/L. In this analysis change in HDL cholesterol was associated with changes in eosinophil and monocyte counts (p<0.001 and p=0.001). An annual increase of 1 mmol/L in HDL cholesterol was associated with an annual decrease in eosinophil and monocytes counts of 0.036×10^9^ and 0.060×10^9^ cells/L, respectively.

### Association of Triglycerides with leukocyte counts

In the cross-sectional analysis (Figure 2, Supplementary Table 2), triglycerides were positively associated with all leukocyte count parameters (all p<0.001). Each one-SD increment in ln-transformed triglycerides (0.52 ln-transformed mmol/L unit) was associated with 3.4%, 0.12%, 3.1%, 2.6%, 3.4% and 4.2% higher total leukocyte, basophil, eosinophil, monocyte, neutrophil and lymphocyte counts respectively. An interaction with sex was found for the association of triglycerides with total leukocyte, eosinophil, monocyte and lymphocyte counts (all p<0.002), with the association being more prominent in women compared to men (Supplementary Table 3).

In the longitudinal analysis (Figure 3, Supplementary Table 4), the average change in triglycerides over the follow up was 0.00±0.83 mmol/L. In this analysis change in triglycerides remained closely associated with changes in total leukocyte, basophil, eosinophil, monocyte and neutrophil counts (all p<0.003). An annual increase of 1 mmol/L in triglycerides was associated with an annual increase in total leukocyte, basophil, eosinophil, monocyte and neutrophil counts of 0.228×10^9^, 0.004×10^9^, 0.015×10^9^, 0.032 ×10^9^ and 0.135×10^9^ cells/L, respectively.

### Association of apoA-I with WBC counts

In the cross-sectional analysis (Figure 2, Supplementary Table 2), apoA-I level was inversely associated with total leukocyte, eosinophil, monocyte and lymphocyte counts (all p<0.001). Each one-SD increment in apoA-I level (0.27 g/L) was associated with 0.5%, 3.0%, 0.9% and 0.8% lower total leukocyte, eosinophil, monocyte and lymphocyte counts, respectively. An interaction with sex was found in the association of apoA-I level with total leukocyte, monocyte and lymphocyte counts (all p<0.01), in which the association with total leukocyte and monocyte counts was more pronounced in women, whereas the association with lymphocyte count was more prominent in men. Although there was no evidence for an association of apoA-I with basophil and neutrophil counts, an interaction with sex was observed (both p<0.001); whereby the association with basophil count was significant in men only and the association with neutrophil count was positive in men yet inverse in women (Supplementary Table 3).

In the longitudinal analysis (Figure 3, Supplementary Table 4), the average change in apoA-I level over the follow up period was 0.05±0.18 g/L. In this analysis change in apoA-I levels demonstrated a significant association with change in eosinophil counts (p<0.001). An annual increase of 1 g/L in apoA-I was associated with an annual decrease in eosinophil count of 0.026×10^9^ cells/L.

### Association of apoB with WBC counts

In cross-sectional analysis (Figure 2, Supplementary Table 2), apoB level was inversely associated with total leukocyte, basophil, eosinophil, monocyte and neutrophil counts (all p<0.001). Each one-SD increment in apoB level (0.24 g/L) was associated with 1.9%, 0.9%, 2.1%, 2.8% and 2.8% lower total leukocyte, basophil, eosinophil, monocyte and neutrophil counts, respectively. A modest, positive association was found between apoB and lymphocyte count; whereby a one-SD increment in apoB level was associated with a 1% increase in lymphocyte count (p=0.011). For the association of apoB level with total leukocyte, monocyte and neutrophil counts, there was a significant interaction with sex (all p<0.01), in which the association was more pronounced in women compared to men. An interaction with sex was also detected for the association of apoB level with eosinophil count (p=0.014), although sex difference in the association was very modest (Supplementary Table 3).

In the longitudinal analysis (Figure 3, Supplementary Table 4), the average change in ApoB level over the follow up period was 0.00±0.19 g/L. In this analysis change in apoB levels showed a strong association with change in lymphocyte counts (p<0.001). An annual increase of 1 g/L in apoB levels was associated with an annual increase in lymphocyte count of 0.320×10^9^ cells/L.

### Association of Lp(a) with WBC counts

In cross-sectional analysis (Figure 2, Supplementary Table 2), Lp(a) level was inversely associated with total leukocyte, monocyte and neutrophil counts (all p<0.001). Each one-SD increment in ln-transformed Lp(a) level (1.12 ln-transformed nmol/L unit) was associated with 0.3%, 0.5% and 0.5% lower total leukocyte, monocyte and neutrophil counts. An interaction with sex was found in the association of Lp(a) level with total leukocyte (p=0.023) and lymphocyte (p=0.026) counts (Supplementary Table 3), in which the association with lower total leukocyte count was more pronounced in females; whereas, the association with higher Lp(a) levels with higher lymphocyte count was more pronounced in males.

In the longitudinal analysis (Figure 3, Supplementary Table 4), the average change in Lp(a) level over the follow up period was 2.6±14.7 nmol/L. In this analysis change in Lp(a) levels was exiguously associated with change in basophil counts, although the association did not reach statistical significance after multiple testing correction. An interaction of sex was detected in this analysis (p=0.001), whereby the association of change in Lp(a) with change in basophil count was apparent in men but not women (Supplementary Table 5).

## Discussion

The cross-sectional relationship of lipids and lipoproteins with leukocyte counts has been previously reported^14-16^. However, such studies were limited by a relatively small sample size and included only basic lipid parameters. By utilising data from the UK Biobank, the present study is the largest to date to assess the cross-sectional relationship between serum lipids and leukocyte count. Further, the data included measures of additional apolipoproteins such as apoA-I, apoB as well as Lp(a), that are involved in the pathophysiology of atherosclerosis. This data was used to analyse the relationship between apoA-I, apoB and Lp(a) and leukocyte counts; a topic where little evidence currently exists. In the cross-sectional analysis, our findings fundamentally align with results from the MESA^15, 16^. In the MESA^16^, NHANES^15^ and UK Biobank studies triglycerides demonstrated the strongest positive association with leukocyte count. The MESA revealed a significant positive association of triglycerides with lymphocyte and total leukocyte counts, whereas the present study yielded a robust positive correlation of triglycerides with all differential leukocyte counts in cross-sectional analysis and all but lymphocytes in longitudinal analysis. Despite controversy as to their precise role in the pathogenesis of atherosclerosis, elevated serum triglycerides are an independent predictor of CVD^20^. Importantly, hypertriglyceridemia is closely linked with inflammation via the generation of free fatty acids released from triglycerides^21^. Free fatty acids promote inflammation by activating NF-κB signalling in mononuclear cells and enhancing reactive oxygen species generation in both mononuclear and polymorphonuclear cells^22^. More recently, free fatty acids have been shown to directly influence myelopoiesis in the bone marrow via their release from bone marrow adipocytes^23^. Given the robust relationship of triglycerides with leukocyte counts in this and previous studies the data indicates that triglycerides may be directly involved in leukogenesis, although to what extent is currently unclear.

ApoA-I, the primary lipoprotein component of HDL has known anti-atherogenic properties mediated largely via its role in cholesterol efflux from cells^24^. Tani *et al*. (2016) previously demonstrated a significant negative correlation of apoA-I concentration with neutrophil, monocyte, eosinophil and total leukocyte counts^25^. Results from this study corroborate the cross-sectional, inverse relationship of apoA-I level with monocyte, eosinophil and total leukocyte counts^25^. In contrast to the earlier report, the results from the present study did not find a significant relationship of apoA-1 levels with neutrophil counts but did identify an inverse association of apoA-I with lymphocyte count. Interestingly, in longitudinal analysis only the relationship apoA-I with eosinophil count remained significant. Although the precise link between apoA-I and eosinophil count remains unclear, apoA-I is known to slow the progression of asthma in mice, at least in part by reducing pulmonary eosinophil infiltration most likely via a reduction in interlukin-5^26^. Thus, it is ostensible that a similar mechanism could occur systemically.

ApoB is the main apolipoprotein constituent of atherogenic lipoproteins including chylomicrons, very-low density lipoproteins, intermediate-density lipoproteins and LDL^27^. In patients with colorectal cancer, serum apoB levels are negatively correlated with neutrophil count^28^ – a finding supported by the cross-sectional analysis in the present study. However, in longitudinal analysis only the association of apoB with lymphocyte remained significant. In previous studies apoB level has been associated with C-reactive protein and monocyte chemoattractant protein-1, suggesting apoB may be a marker of inflammation^28, 29^. Whether apoB is directly involved in promoting inflammation or whether it is a biproduct of the inflammatory cascade remains unclear.

Serum Lp(a) levels are an independent predictor of CVD and mendelian randomization studies have demonstrated a direct role of Lp(a) in the pathogenesis of atherosclerosis^30, 31^. To our knowledge, no published literature has assessed the association of Lp(a) with leukocyte counts. We observed a modest negative association of Lp(a) level with monocyte, neutrophil and total leukocyte count in cross-sectional analysis. However, as the level of Lp(a) is predominantly genetically determined only a small, likely serendipitous change in Lp(a) concentration was observed over the follow-up period^32^. As such, this data suggests that the role of Lp(a) in CVD is independent of any effects on leukogenesis.

Although the cross-sectional relationship of lipids and lipoproteins with leukocyte counts provides an important snapshot into the pathobiology of CVD, it does not take into account the chronic nature of this disease. CVD develops over decades, with atheroma formation beginning as early as the second decade of life in apparently heathy individuals^33^. As such it important to assess not only cross-sectional but also longitudinal relationships^34^. Overall, many of the associations seen in cross-sectional analysis did not remain significant in the longitudinal analysis. A likely explanation for this is the relatively short follow-up period resulting in little change of lipid, lipoprotein and leukocyte counts over this time. Also, the very large sample size in the cross-sectional analysis allowed detection of minute changes, which may not be clinically significant. Therefore, longitudinal analysis was used to identify the most robust relationships. Beyond this longitudinal analysis provides important insight into how these parameters interact over time. However, due to the lack of biochemical data at subsequent follow-up visits it was not possible to assess the temporality of these relationships. As the UK Biobank matures this data may become available and allow further characterisation of the precise relationship of lipids and lipoproteins with leukocyte counts. Additionally, as this was an observational study, further preclinical research is required to delineate the underlying mechanism of these associations.

Overall, by utilising a large, comprehensive dataset this study highlights significant cross-sectional and longitudinal relationships of lipid and lipoprotein parameters with leukocyte counts. However, unlike in murine models the majority of these relationships are modest, with the most robust association being between triglycerides and differential leukocyte counts. This provides further evidence that major differences exist between murine and human models of atherosclerosis, particularly with respect to the interaction between lipids, lipoproteins and haematopoiesis.

## Data Availability

All data is available from the authors by request

## Acknowledgements

This research was conducted using the UK Biobank resource. The authors would like to acknowledge all study participants and personnel who make this resource possible.

## References

1. Libby P. Inflammation in Atherosclerosis. Arteriosclerosis, Thrombosis, and Vascular Biology 2012;32(9):2045–2051.

2. Bentzon Jacob F, Otsuka F, Virmani R, Falk E. Mechanisms of Plaque Formation and Rupture. Circulation Research 2014;114(12):1852–1866.

3. Rye KA, Bursill CA, Lambert G, Tabet F, Barter PJ. The metabolism and anti-atherogenic properties of HDL. J Lipid Res 2009;50 Suppl:S195–200.

4. Weber C, Noels H. Atherosclerosis: current pathogenesis and therapeutic options. Nature Medicine 2011;17(11):1410–1422.

5. Nordestgaard BG, Chapman MJ, Ray K, Borén J, Andreotti F, Watts GF, Ginsberg H, Amarenco P, Catapano A, Descamps OS, Fisher E, Kovanen PT, Kuivenhoven JA, Lesnik P, Masana L, Reiner Z, Taskinen M-R, Tokgözoglu L, Tybjærg-Hansen A, European Atherosclerosis Society Consensus P. Lipoprotein(a) as a cardiovascular risk factor: current status. European heart journal 2010;31(23):2844–2853.

6. Patel A, Barzi F, Jamrozik K, Lam TH, Ueshima H, Whitlock G, Woodward M. Serum triglycerides as a risk factor for cardiovascular diseases in the Asia-Pacific region. Circulation 2004;110(17):2678–86.

7. Sniderman Allan D, Williams K, Contois John H, Monroe Howard M, McQueen Matthew J, de Graaf J, Furberg Curt D. A Meta-Analysis of Low-Density Lipoprotein Cholesterol, Non-High-Density Lipoprotein Cholesterol, and Apolipoprotein B as Markers of Cardiovascular Risk. Circulation: Cardiovascular Quality and Outcomes 2011;4(3):337–345.

8. Dawson TC, Kuziel WA, Osahar TA, Maeda N. Absence of CC chemokine receptor-2 reduces atherosclerosis in apolipoprotein E-deficient mice. Atherosclerosis 1999;143(1):205–11.

9. Welsh C, Welsh P, Mark PB, Celis-Morales CA, Lewsey J, Gray SR, Lyall DM, Iliodromiti S, Gill JMR, Pell J, Jhund PS, Sattar N. Association of Total and Differential Leukocyte Counts With Cardiovascular Disease and Mortality in the UK Biobank. Arterioscler Thromb Vasc Biol 2018;38(6):1415–1423.

10. Soehnlein O, Swirski FK. Hypercholesterolemia links hematopoiesis with atherosclerosis. Trends in endocrinology and metabolism: TEM 2013;24(3):129–136.

11. Gomes AL, Carvalho T, Serpa J, Torre C, Dias S. Hypercholesterolemia promotes bone marrow cell mobilization by perturbing the SDF-1:CXCR4 axis. Blood 2010;115(19):3886–94.

12. Karpova D, Bonig H. Concise Review: CXCR4/CXCL12 Signaling in Immature Hematopoiesis—Lessons From Pharmacological and Genetic Models. STEM CELLS 2015;33(8):2391–2399.

13. Murphy AJ, Akhtari M, Tolani S, Pagler T, Bijl N, Kuo CL, Wang M, Sanson M, Abramowicz S, Welch C, Bochem AE, Kuivenhoven JA, Yvan-Charvet L, Tall AR. ApoE regulates hematopoietic stem cell proliferation, monocytosis, and monocyte accumulation in atherosclerotic lesions in mice. J Clin Invest 2011;121(10):4138–49.

14. Liu Y, Kong X, Wang W, Fan F, Zhang Y, Zhao M, Wang Y, Wang Y, Wang Y, Qin X, Tang G, Wang B, Xu X, Hou FF, Gao W, Sun N, Li J, Venners SA, Jiang S, Huo Y. Association of peripheral differential leukocyte counts with dyslipidemia risk in Chinese patients with hypertension: insight from the China Stroke Primary Prevention Trial. Journal of lipid research 2017;58(1):256–266.

15. Andersen CJ, Vance TM. Gender Dictates the Relationship between Serum Lipids and Leukocyte Counts in the National Health and Nutrition Examination Survey 1999L2004. Journal of clinical medicine 2019;8(3):365.

16. Lai YC, Woollard KJ, McClelland RL, Allison MA, Rye KA, Ong KL, Cochran BJ. The association of plasma lipids with white blood cell counts: Results from the Multi-Ethnic Study of Atherosclerosis. J Clin Lipidol 2019;13(5):812–820.

17. Elliott P, Peakman TC. The UK Biobank sample handling and storage protocol for the collection, processing and archiving of human blood and urine. Int J Epidemiol 2008;37(2):234–44.

18. Sudlow C, Gallacher J, Allen N, Beral V, Burton P, Danesh J, Downey P, Elliott P, Green J, Landray M, Liu B, Matthews P, Ong G, Pell J, Silman A, Young A, Sprosen T, Peakman T, Collins R. UK biobank: an open access resource for identifying the causes of a wide range of complex diseases of middle and old age. PLoS medicine 2015;12(3):e1001779–e1001779.

19. Townsend P. Deprivation. Journal of Social Policy 1987;16(2):125–146.

20. Miller M, Stone Neil J, Ballantyne C, Bittner V, Criqui Michael H, Ginsberg Henry N, Goldberg Anne C, Howard William J, Jacobson Marc S, Kris-Etherton Penny M, Lennie Terry A, Levi M, Mazzone T, Pennathur S. Triglycerides and Cardiovascular Disease. Circulation 2011;123(20):2292–2333.

21. Tenenbaum A, Klempfner R, Fisman EZ. Hypertriglyceridemia: a too long unfairly neglected major cardiovascular risk factor. Cardiovascular Diabetology 2014;13(1):159.

22. Tripathy D, Mohanty P, Dhindsa S, Syed T, Ghanim H, Aljada A, Dandona P. Elevation of Free Fatty Acids Induces Inflammation and Impairs Vascular Reactivity in Healthy Subjects. Diabetes 2003;52(12):2882.

23. Robles H, Park S, Joens MS, Fitzpatrick JAJ, Craft CS, Scheller EL. Characterization of the bone marrow adipocyte niche with three-dimensional electron microscopy. Bone 2019;118:89–98.

24. Rye K-A. High density lipoprotein structure, function, and metabolism: a new Thematic Series. Journal of Lipid Research 2013;54(8):2031–2033.

25. Tani S, Nagao K, Hirayama A. Association of systemic inflammation with the serum apolipoprotein A-1 level: A cross-sectional pilot study. Journal of Cardiology 2016;68(2):168–177.

26. Yao X, Gordon EM, Figueroa DM, Barochia AV, Levine SJ. Emerging Roles of Apolipoprotein E and Apolipoprotein A-I in the Pathogenesis and Treatment of Lung Disease. American journal of respiratory cell and molecular biology 2016;55(2):159–169.

27. Shapiro MD, Fazio S. Apolipoprotein B-containing lipoproteins and atherosclerotic cardiovascular disease. F1000Research 2017;6:134–134.

28. Sirniö P, Väyrynen JP, Klintrup K, Mäkelä J, Mäkinen MJ, Karttunen TJ, Tuomisto A. Decreased serum apolipoprotein A1 levels are associated with poor survival and systemic inflammatory response in colorectal cancer. Scientific Reports 2017;7(1):5374.

29. Xu W, Li R, Zhang S, Gong L, Wang Z, Ren W, Xia C, Li Q. The relationship between high-sensitivity C-reactive protein and ApoB, ApoB/ApoA1 ratio in general population of China. Endocrine 2012;42(1):132–8.

30. Cegla J, Neely RDG, France M, Ferns G, Byrne CD, Halcox J, Datta D, Capps N, Shoulders C, Qureshi N, Rees A, Main L, Cramb R, Viljoen A, Payne J, Soran H. HEART UK consensus statement on Lipoprotein(a): A call to action. Atherosclerosis 2019;291:62–70.

31. Saleheen D, Haycock PC, Zhao W, Rasheed A, Taleb A, Imran A, Abbas S, Majeed F, Akhtar S, Qamar N, Zaman KS, Yaqoob Z, Saghir T, Rizvi SNH, Memon A, Mallick NH, Ishaq M, Rasheed SZ, Memon FU, Mahmood K, Ahmed N, Frossard P, Tsimikas S, Witztum JL, Marcovina S, Sandhu M, Rader DJ, Danesh J. Apolipoprotein(a) isoform size, lipoprotein(a) concentration, and coronary artery disease: a mendelian randomisation analysis. Lancet Diabetes Endocrinol 2017;5(7):524–533.

32. McCormick SPA. Lipoprotein(a): biology and clinical importance. The Clinical biochemist. Reviews 2004;25(1):69–80.

33. Insull W, Jr. The pathology of atherosclerosis: plaque development and plaque responses to medical treatment. Am J Med 2009;122(1 Suppl):S3–s14.

34. Caruana EJ, Roman M, Hernández-Sánchez J, Solli P. Longitudinal studies. Journal of thoracic disease 2015;7(11):E537–E540.

